# Improving Prediction of Patient-Reported Health Outcomes Among Older Adults With Diabetes Across Diverse Populations Through Transfer Learning

**DOI:** 10.64898/2026.11.24.26368865

**Authors:** Xuzhi Wang, Yuanyuan Chen, Weipeng Shao, Jeannette M. Beasley, Ting-Fang Lee

## Abstract

Population heterogeneity challenges the transportability of prediction models. Transfer learning may improve prediction through information borrowing, but its utility across population settings remains unclear. We evaluated a regression-based transfer learning framework across international aging cohorts (Gateway) and race/ethnicity groups within the RAND Health and Retirement Study (RAND HRS). Gateway included 19,879 adults with diabetes from eight cohorts, while RAND HRS included 3,926 participants across four race/ethnicity groups. Prediction models for patient-reported health outcomes were developed using demographic and clinical risk factors. Transfer learning was compared with target-only and pooled models using AUC and Brier score, with bootstrap resampling to assess uncertainty. Transfer learning improved prediction in 75% (6/8) of Gateway cohorts, with AUC gains of 0.1%–0.6%, and in all four RAND HRS race/ethnicity groups, with gains of 0.8%–3.1%. Improvements were greater and more consistent across within-cohort groups. These findings demonstrate the potential of regression-based transfer learning to improve prediction across heterogeneous populations. Information borrowing from sufficiently transferable external data may enhance prediction performance and provide meaningful public health benefits, even when improvements are modest, particularly in large population-based cohorts.

## Introduction

Diabetes is a major and growing global public health challenge. In 2024, an estimated 589 million adults aged 20–79 years were living with diabetes worldwide, and this number is projected to increase to 853 million by 2050 ^[1]^. Diabetes is highly prevalent among older adults, and continued population aging is expected to further increase the number of older adults living with the disease ^[1]^. Beyond major clinical complications, people with diabetes may experience a range of health problems affecting functioning, including pain and poor visual health ^[2]^. These aspects of health can be captured through patient-reported outcomes, such as self-reported eyesight, which provide information complementary to conventional clinical measures ^[3]^.

Numerous prediction models have been developed for health outcomes among people with diabetes, using approaches ranging from conventional statistical methods to more recent machine-learning techniques ^[4,5]^. Many have shown promising predictive performance in their development populations ^[5]^. However, prediction models are often developed within specific populations and healthcare settings, which may limit their applicability to other settings ^[6,7]^. External validation is therefore important for assessing model generalizability and transportability across populations and settings ^[7,8]^.

Differences in population characteristics and healthcare settings can limit the performance of prediction models when they are applied beyond the populations in which they were developed ^[9,10]^. Populations may differ in their composition and healthcare context, resulting in differences in case mix and data distributions across settings ^[7,10]^. Such heterogeneity can contribute to differences in predictor–outcome relationships and variation in predictive performance across populations ^[11,12]^. Indeed, external validation studies have documented variation in predictive performance across populations and settings ^[13,14]^.

One approach to addressing population heterogeneity is to develop prediction models separately within each target population. Although this approach allows models to capture population-specific characteristics, reliable model development requires sufficiently large, high-quality target data, which may not be readily available for all populations ^[15,16]^. Alternatively, pooling data across populations can increase the available sample size but may fail to adequately account for differences in predictor–outcome relationships across populations. These limitations motivate approaches that leverage information from related populations while accounting for population-specific differences.

Transfer learning has emerged as a promising approach for leveraging information from related populations while accommodating differences between source and target populations ^[17]^. Rather than assuming that source and target populations are identical, transfer learning seeks to exploit information shared across related populations while accounting for differences between them ^[18]^. The regression-based transfer learning framework applied in this study first combines information from the source and target populations to obtain an initial model, thereby leveraging the additional information available from the source data. The model is then refined using target-population data to correct for differences between the source and target populations ^[18]^. When the differences between source and target models are relatively limited, borrowing source information can reduce estimation error compared with relying on target data alone ^[18]^. This balance between information sharing and population specificity makes regression-based transfer learning particularly appealing for prediction across heterogeneous populations.

Despite its potential, the effectiveness of transfer learning depends on the relationship between source and target populations. Theoretical work has shown that transfer learning can improve estimation and prediction when source and target populations are sufficiently similar, whereas borrowing information across populations with greater differences may provide limited benefits or even adversely affect predictive performance ^[18,19]^. However, how these theoretical notions of source-target similarity translate to the complex heterogeneity observed across real-world populations remains less well understood. Although population similarity is generally expected to favor information transfer, there remains limited empirical evidence regarding when differences between populations become sufficiently substantial to diminish the benefits of information borrowing ^[20,21]^. A better understanding of these conditions is important for determining when information borrowing can be effectively used to improve prediction across diverse populations.

In this study, we evaluated a regression-based transfer learning framework for predicting patient-reported health outcomes among adults with diabetes across two distinct population settings. We examined between-country differences using harmonized international aging cohorts and within-country differences across racial and ethnic groups in the RAND Health and Retirement Study (RAND HRS). The outcome was defined by the presence of pain or visual impairment in the international cohorts and by self-reported overall eyesight in the RAND HRS. In both applications, we compared transfer learning with target-only and pooled models. These comparisons were used to assess the predictive benefit of borrowing information from related populations.

## Methods

### Study Design and Data Sources

We conducted two complementary analyzes representing distinct forms of population heterogeneity: an across-cohort analysis and a within-cohort analysis. Both analyzes used the same regression-based transfer learning framework (Figure 1) and the model evaluation strategy, but differed in data preparation, outcome definition, and population structure.

**Figure 1.**
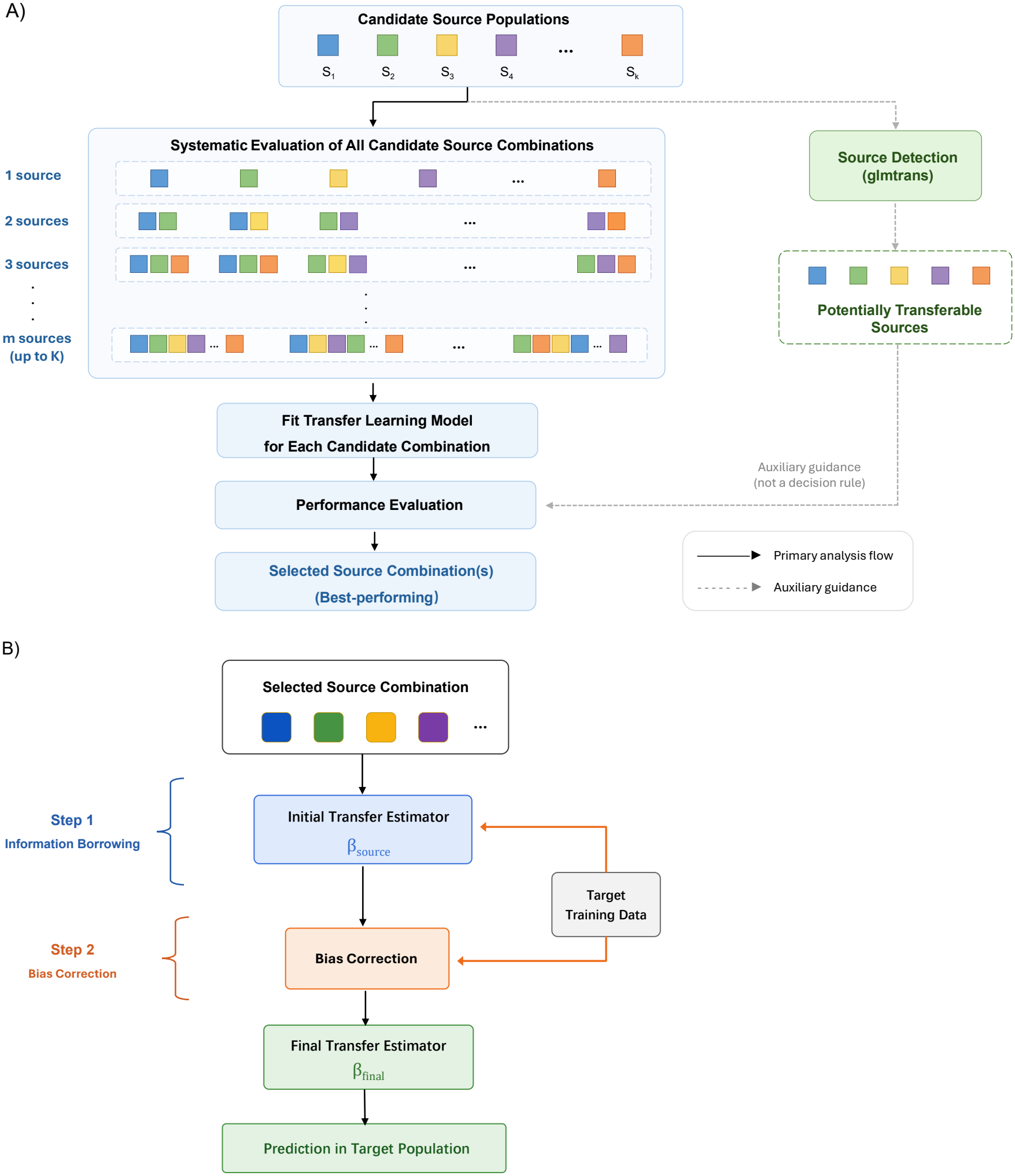
Overview of the proposed regression-based transfer learning framework. (A) Systematic evaluation of candidate source combinations. (B) Two-step transfer learning framework with information borrowing and bias correction.

The across-cohort analysis used harmonized data from eight prospective longitudinal aging cohorts available through the Gateway to Global Aging Data platform ^[22]^, including the Health and Retirement Study (HRS, United States) ^[23]^, the Mexican Health and Aging Study (MHAS, Mexico) ^[24]^, the Longitudinal Aging Study in India (LASI, India) ^[25]^, the China Health and Retirement Longitudinal Study (CHARLS, China) ^[26]^, the Survey of Health, Ageing and Retirement in Europe (SHARE, Europe) ^[27]^, the English Longitudinal Study of Ageing (ELSA, United Kingdom) ^[28]^, the Korean Longitudinal Study of Aging (KLoSA, South Korea) ^[29]^, and the Health and Aging in Africa: A Longitudinal Study of an INDEPTH Community in South Africa (HAALSI, South Africa) ^[30]^. For each cohort, the survey wave closest to 2018 was selected to improve temporal comparability across datasets. The analytic population was restricted to participants aged 55 years or older with diagnosed diabetes; cohort-specific distributions of diabetes status before participant selection are provided in Supplementary Table S1. For the within-cohort analysis, data were obtained from the RAND HRS Longitudinal File, with analyses restricted to the 2018 wave ^[31,32]^. The HRS was selected because its racial/ethnic diversity allows the evaluation of transfer learning across subpopulations within a single country. Additionally, using data from a single cohort eliminates the need for cross-country variable harmonization, allowing the analysis to focus on population heterogeneity without differences in variable definitions across cohorts.

### Data Preparation and Variable Construction

Different data preparation procedures were implemented for the Gateway and HRS applications because they addressed distinct population heterogeneity settings.

For the across-cohort application, differences in questionnaire design, healthcare systems, and variable definitions across cohorts were addressed using the Gateway harmonization framework ^[22]^. Non-substantive survey responses, including “Don’t know,” “Refused,” and “Not ascertained,” were recoded as missing values rather than treated as analytic categories, and missingness arising from cohort-specific questionnaire logic or skip patterns was distinguished from item non-response ^[22]^.

The harmonized predictor set included variables across demographic, socioeconomic, behavioral, and clinical domains (Table 1), with further details on variable definitions and harmonization procedures provided in Supplementary Table S2. To improve across-cohort comparability, the socioeconomic indicator was converted to international dollars using country-specific 2018 Purchasing Power Parity (PPP) conversion factors from the World Bank ^[33]^. Missing predictor values were imputed separately within each cohort using Multiple Imputation by Chained Equations (MICE) under the Missing at Random (MAR) assumption ^[34]^. For the SHARE cohort, selected predictor variables exhibited substantial missingness (Supplementary Table S3); therefore, continuous variables were imputed using cohort-level means and categorical variables using cohort-level modes.

**Table 1.** Baseline characteristics of participants with diabetes across eight aging cohorts.

| Characteristic | Statistic | HRS<br>(*n* = 4449) | MHAS<br>(*n* = 3488) | LASI<br>(*n* = 6301) | CHARLS<br>(*n* = 2127) | ELSA<br>(*n* = 269) | KLoSA<br>(*n* = 1382) | HAALSI<br>(*n* = 509) | SHARE<br>(*n* = 10 367) |
|---|---|---|---|---|---|---|---|---|---|
| **Outcome characteristics** |  |  |  |  |  |  |  |  |  |
| Visual impairment | n (%) | 577 (13.0) | 351 (10.1) | 1577 (25.0) | 702 (33.0) | 6 (2.2) | 90 (6.5) | 164 (32.2) | 955 (1.2) |
| Presence of pain | n (%) | 2136 (48.0) | 1474 (42.3) | 3315 (52.6) | 1442 (67.8) | 115 (42.8) | 577 (41.8) | 161 (31.6) | 1248 (12.0) |
| Kidney disease | n (%) | NA | 141 (4.0) | NA | 340 (16.0) | NA | NA | 11 (2.2) | 498 (4.8) |
| **Demographic characteristics** |  |  |  |  |  |  |  |  |  |
| Female sex | n (%) | 2535 (57.0) | 2140 (61.4) | 2084 (33.1) | 1176 (55.3) | 128 (47.6) | 774 (56.0) | 299 (58.7) | 5438 (52.5) |
| Age, years | Mean (SD) | 69.9 (9.8) | 69.0 (26.6) | 66.0 (7.7) | 66.6 (8.2) | 69.9 (9.3) | 73.1 (9.0) | 70.3 (10.0) | 71.8 (8.7) |
|  | Median (IQR) | 68 (61–77) | 68 (61–74) | 65 (60–71) | 65 (60–72) | 70 (63–77) | 73 (66–80) | 69 (62–76) | 71 (65–78) |
| **Behavioral characteristics** |  |  |  |  |  |  |  |  |  |
| Ever smoked | n (%) | 2498 (56.2) | 1245 (35.7) | 1681 (26.7) | 841 (39.5) | 165 (61.3) | 107 (7.7) | 52 (10.2) | 29 (0.28) |
| Former smoker | n (%) | 2032 (45.7) | 972 (27.9) | 863 (13.7) | 330 (15.5) | 137 (50.9) | NA | 32 (6.3) | NA |
| Adherence to diabetic diet | n (%) | NA | 1861 (53.4) | NA | NA | 17 (6.3) | NA | 56 (11.0) | NA |
| **Clinical characteristics** |  |  |  |  |  |  |  |  |  |
| Receiving diabetes treatment | n (%) | 2992 (67.3) | 3242 (93.0) | 5253 (83.4) | 675 (31.7) | 127 (47.2) | 1154 (83.5) | 162 (31.8) | 9150 (88.3) |
| Use of antihypertensive medication | n (%) | 3321 (74.6) | 2162 (62.0) | 3569 (56.6) | 1082 (50.9) | 128 (47.6) | 1246 (90.2) | 261 (51.3) | 7625 (73.6) |
| **Socioeconomic characteristics** |  |  |  |  |  |  |  |  |  |
| Public health insurance | n (%) | 3326 (74.8) | 1857 (53.2) | 1386 (22.0) | 2078 (97.7) | NA | 1382 (100.0) | 62 (12.2) | 1027 (9.9) |
| Pension income | Mean (SD) | 23 683.4 (82 666.8) | 379.0 (595.5) | 546.2 (1506.0) | 3548.5 (4595.1) | 77.8 (115.8) | 336.4 (552.1) | 1774.4 (823.2) | 2342.4 (22 132.8) |
|  | Median (IQR) | 13 572 (5022–28 629) | 215.6 (118.6–404.7) | 0 (0–195.4) | 488.5 (244.3–6350.6) | 27.7 (0–122.2) | 241 (0–373) | 1700 (1700–1700) | 0 (0–0) |
| Social assistance benefits | Mean (SD) | 14 293.0 (11 681.5) | 62.8 (73.3) | 72.2 (269.4) | 879.0 (3223.2) | 18.9 (39.0) | 688.8 (183.3) | 1022.4 (714.1) | 66.8 (1553.5) |
|  | Median (IQR) | 13 200 (9000–18 000) | 53.9 (26.7–62.5) | 7.3 (0–46.9) | 256.5 (129.5–586.2) | 0 (0–17.5) | 673 (553–841) | 800 (410–1230) | 0 (0–0) |

For the within-cohort application, the same categories of predictor variables (Table 2) were derived directly from the RAND HRS Longitudinal File without cross-cohort harmonization. Because missingness was minimal across all model variables (all below 3%), complete-case analysis was adopted, with participants excluded if any predictor or outcome variable was missing.

**Table 2.** Baseline characteristics of participants with diabetes in the RAND Health and Retirement Study (RAND HRS).

| Characteristic | Statistic | RAND HRS<br>(*n* = 4259) |
|---|---|---|
| **Outcome characteristics** |  |  |
| Self-reported eyesight | Poor | 375 (8.8) |
|  | Fair | 1007 (23.6) |
|  | Good | 1795 (42.1) |
|  | Very good | 790 (18.6) |
|  | Excellent | 281 (6.6) |
| **Demographic characteristics** |  |  |
| Female sex | n (%) | 2424 (56.9) |
| Race/ethnicity | Hispanic | 939 (22.0) |
|  | Non-Hispanic White | 2007 (47.1) |
|  | Non-Hispanic Black | 1100 (25.8) |
|  | Other non-Hispanic | 208 (4.9) |
| Age, years | Mean (SD) | 69.6 (9.8) |
|  | Median (IQR) | 68 (61–77) |
| **Behavioral characteristics** |  |  |
| Smoking status | Current | 440 (10.3) |
|  | Former | 1934 (45.4) |
|  | Never | 1869 (43.9) |
| **Clinical characteristics** |  |  |
| Diabetes treatment | No treatment | 1151 (27.0) |
|  | Oral only | 2276 (53.4) |
|  | Insulin only | 25 (0.59) |
|  | Oral + insulin | 686 (16.1) |
| BMI | Mean (SD) | 31.0 (6.7) |
|  | Median (IQR) | 30 (27–34) |
| **Socioeconomic characteristics** |  |  |
| Education | Less than high school | 986 (23.1) |
|  | General Educational Development | 237 (5.6) |
|  | High school graduate | 1165 (27.4) |
|  | Some college | 1064 (25.0) |
|  | College or above | 807 (19.0) |
| Health insurance | Medicare | 2905 (68.2) |
|  | Medicaid | 821 (19.3) |
|  | Private insurance | 1620 (38.0) |
| Wealth | Mean (SD) | 357285 (1001640) |
|  | Median (IQR) | 99000 (4000–333000) |

add the definition of the numbers of wealth (GDP?) in the footnote

### Outcome Definition

Different outcome definitions were adopted for the Gateway and RAND HRS studies because they addressed distinct prediction settings.

For the across-cohort application, because harmonized clinical biomarkers and adjudicated complication registries were not consistently available across the included cohorts, a proxy-based outcome framework was adopted using harmonized survey measures available across datasets ^[22,35]^. Pain-related burden and self-reported visual impairment were selected as proxy indicators because of their clinical and functional relevance among older adults with diabetes ^[2,36]^. Kidney disease metrics were excluded because several cohorts lacked systematic data collection regarding nephropathy, and their inclusion would have introduced structural missingness across cohorts. The two proxy indicators were each coded as binary variables and combined to define the binary outcome. Participants were classified as having diabetes complication burden if one or more proxy indicators were present; otherwise, they were classified as having no diabetes complication burden.

For the within-cohort application, the outcome was self-rated vision impairment derived from the RAND HRS Longitudinal File. Participants were classified as having vision impairment if they reported fair or poor eyesight; otherwise, they were classified as having no vision impairment.

### Transfer Learning Framework

We compared three complementary prediction strategies within a unified regression-based prediction framework: a target-only baseline model, a pooled model, and a regression-based transfer learning model. The overall framework consisted of two sequential components: source selection and transfer learning, as illustrated in Figure 1.

#### Source selection (Figure 1A)

All available source populations were first considered as candidate sources for transfer learning. Candidate source combinations ranging from single-source to multi-source transfer were systematically generated and evaluated under the same transfer learning framework. The source combination yielding the best predictive performance was subsequently selected for model development.

Transferable source detection was additionally performed to assess the transferability of individual source populations ^[18]^. The resulting transferability estimates complemented the systematic evaluation of candidate source combinations during source selection.

#### Transfer learning (Figure 1B)

After the optimal source combination had been identified, regression-based transfer learning was performed using a two-step procedure consisting of information borrowing and bias correction. The mathematical formulation of the two-step procedure is described below.

Let *L*(·) denote the average negative log-likelihood function of a generalized linear model.

### Step 1: Information Borrowing

Instead of estimating model coefficients using only the target training data, transfer learning first borrows information from the selected source combination. The initial transfer estimator is obtained by minimizing the penalized negative log-likelihood over the pooled target (T) and selected source (S) training data:

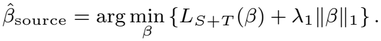

Here, *L*_*S*+*T*_ (·) denotes the average negative log-likelihood evaluated using the pooled target and selected source training data, and *λ*_1_ is the regularization parameter.

### Step 2: Bias Correction

The initial transfer estimator may remain biased because systematic differences can exist between the selected source populations and the target population. Therefore, a bias-correction term is estimated using only the target training data:

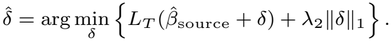

Here, *L*_*T*_ (·) denotes the average negative log-likelihood evaluated using the target training data.

### Output: Final Transfer Estimator

The final transfer estimator is obtained by combining the initial transfer estimator with the estimated bias-correction term:

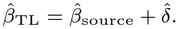

Predictions in the target population were subsequently generated using 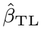.

### Model Development and Training Strategy

After the optimal source combination had been identified, model development was conducted under a unified training strategy to ensure a fair comparison across the three prediction approaches.

For each target population, participants were randomly divided into training (70%) and testing (30%) sets using stratified sampling to preserve the outcome distribution. All selected source populations were retained in their entirety for model development, whereas only the target population was partitioned into training and testing sets. Within each repetition, the same target training and testing split was applied to all three prediction approaches.

The target-only baseline model, pooled model, and regression-based transfer learning model were subsequently implemented under this common training strategy. The pooled model combined the target training data with the selected source population(s), whereas the transfer learning model was implemented using the regression-based framework described in the previous section through the `glmtrans` package ^[37]^. To ensure methodological consistency across prediction approaches, the baseline and pooled models were both fitted using ridge-penalized logistic regression, with the regularization parameter selected by cross-validation ^[38]^.

The entire model development procedure was repeated 300 times using independent random training–testing splits. Model performance from each repetition was retained for subsequent evaluation and statistical analysis.

### Model Evaluation and Statistical Analysis

Predictive performance was evaluated using three measures that capture different aspects of model performance. Discrimination was assessed using the area under the receiver operating characteristic curve (AUC), overall classification performance was evaluated using prediction accuracy, and overall prediction error was assessed using the Brier score, with lower values indicating better predictive performance.

Model performance was summarized across the repeated model development procedures described above. For each prediction strategy, the mean and standard deviation of each performance metric were calculated. Performance gains were quantified as the differences between the transfer learning model and the corresponding comparison model (target-only baseline or pooled model). Corresponding 95% confidence intervals were calculated assuming approximate normality of the repeated performance estimates, and statistical significance was assessed using one-sample *t*-tests on the repeated performance gains.

All analyses were implemented in R version 4.4.2 (R Foundation for Statistical Computing) using the `glmtrans` package and other standard R packages.

## Results

### Participant Characteristics

The Gateway analysis included 28,892 participants with diabetes from eight international aging cohorts (Table 1). Participant characteristics varied substantially across cohorts. Several variables had substantial missingness in SHARE (Supplementary Table S3), and expanded descriptive statistics across cohorts are provided in Supplementary Tables S4–S5.

The HRS analysis included 4,259 participants with diabetes from four racial/ethnic groups (Table 2). Participant characteristics varied across racial/ethnic groups, with additional details provided in Supplementary Table S6.

### Prediction Performance of Transfer Learning

Source detection indicated that the potentially transferable sources differed across target populations. Detailed results across the different population settings are provided in Supplementary Tables S7 and S8. Figure 2 compares the performance of the pooled and transfer learning models relative to the target-only baseline across the international aging cohorts (Gateway) and the Health and Retirement Study (HRS) race/ethnicity groups. In the Gateway analysis, transfer learning improved prediction performance relative to the target-only baseline in 6 of the 8 target cohorts (75%) (Figure 2A). Among these cohorts, gains in AUC ranged from approximately 0.1% to 0.6%. Transfer learning showed no clear improvement over the target-only baseline in the UK cohort, while a small decrease in AUC was observed for SHARE. Compared with the pooled model, transfer learning generally achieved higher AUCs across the Gateway cohorts, with the largest differences observed for CHARLS, SHARE, and LASI. Complete prediction performance results are provided in Supplementary Table S9.

**Figure 2.**
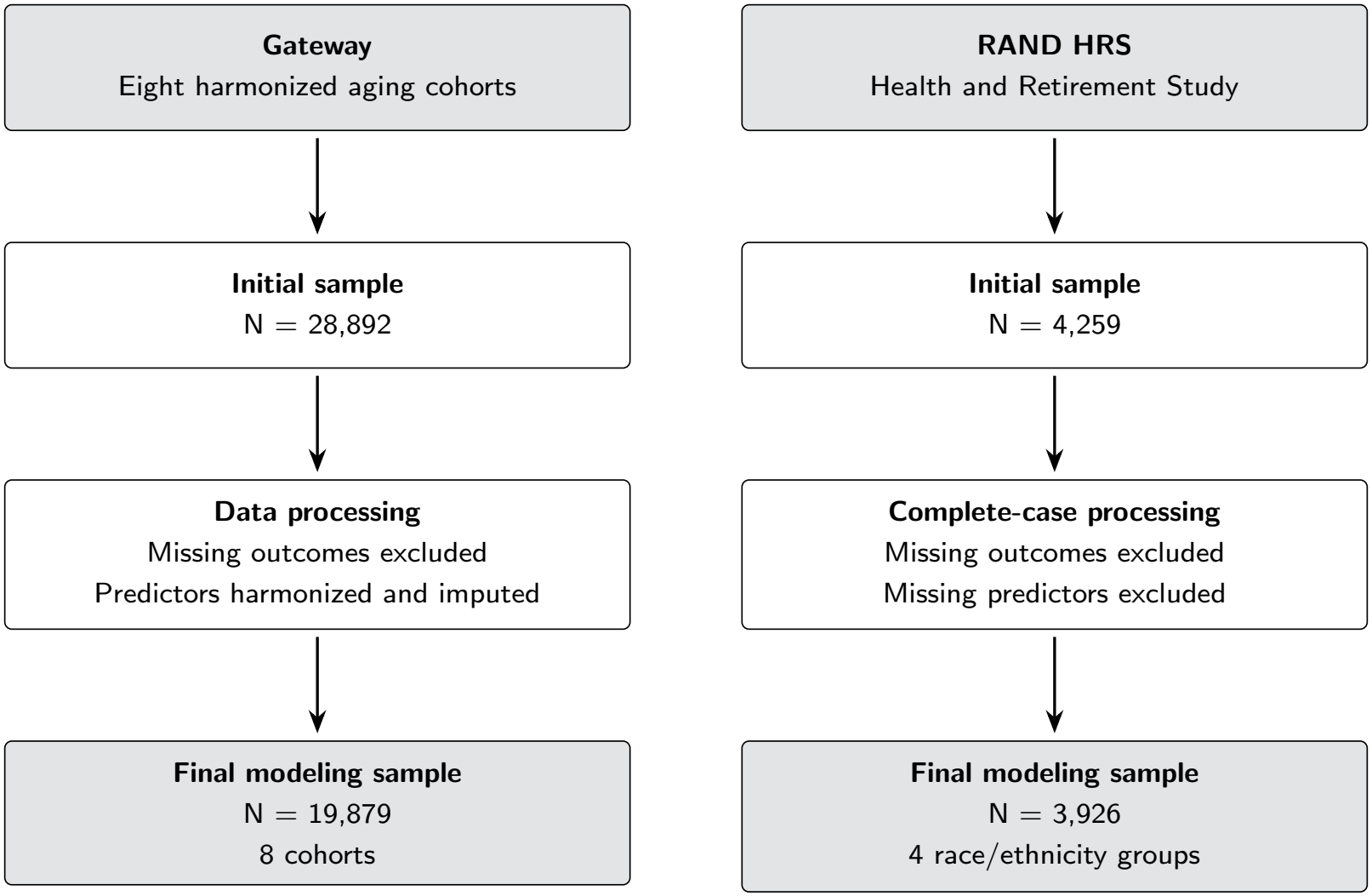
Flowchart of sample construction for the Gateway and RAND HRS analyses, summarizing how the final modeling samples were derived. HRS, Health and Retirement Study.

**Figure 3.**
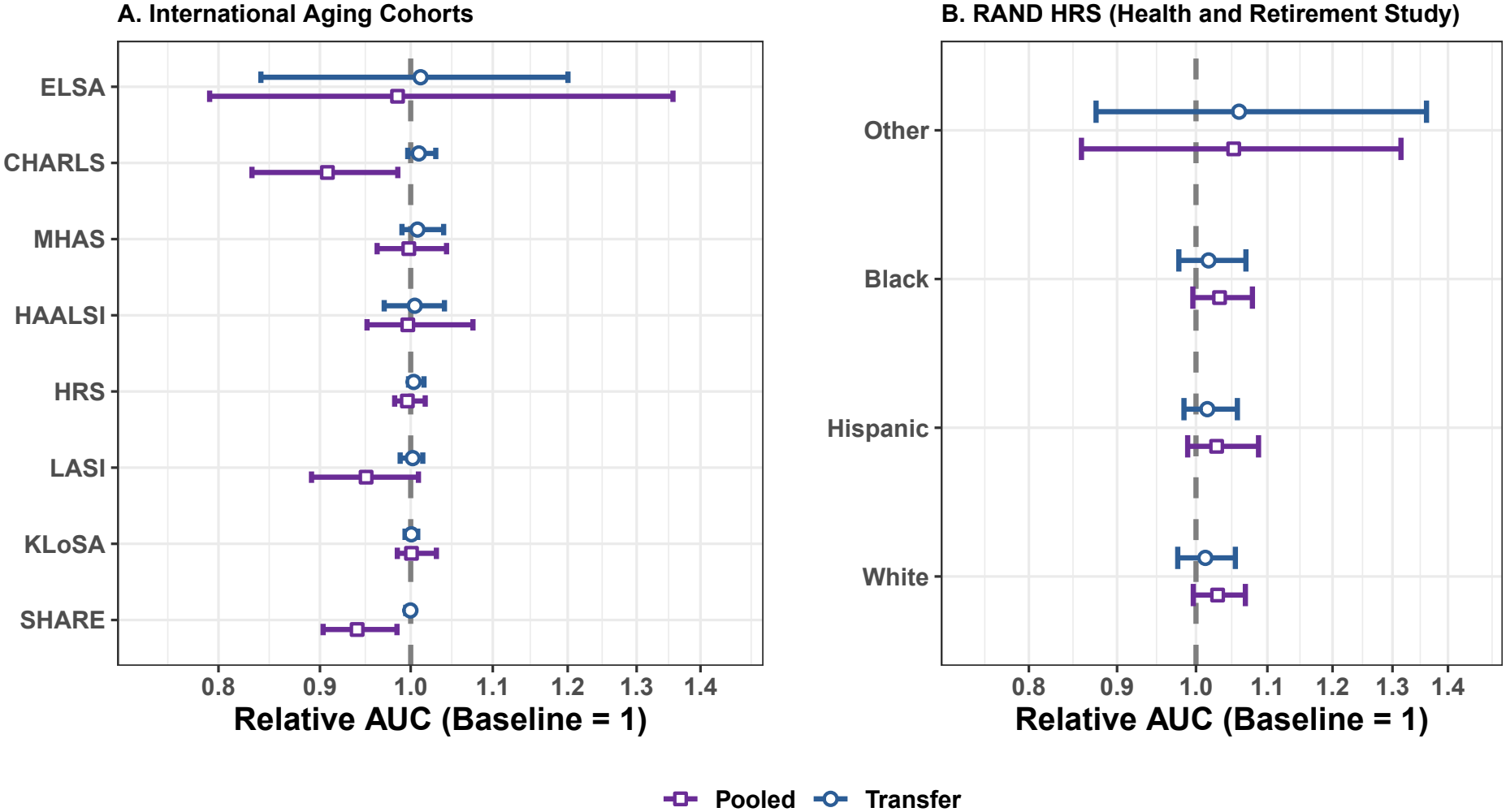
Relative prediction performance of pooled and transfer learning models compared with the target-only baseline model across the International Aging Cohorts (Gateway) and the Health and Retirement Study (HRS) race/ethnicity groups. Relative AUC was expressed as the ratio of the model AUC to the target-only baseline AUC (baseline = 1). Error bars indicate empirical 95% intervals. Detailed performance results are provided in Supplementary Tables S9 and S10.

In the HRS analysis, transfer learning improved prediction performance relative to the target-only baseline across all four race/ethnicity groups (100%) (Figure 2B). AUC gains ranged from approximately 0.8% among non-Hispanic White participants to 3.1% among other non-Hispanic participants. The pooled model also improved prediction relative to the target-only baseline across all four groups and achieved higher AUCs than transfer learning among non-Hispanic Black, Hispanic, and non-Hispanic White participants, whereas transfer learning achieved the highest AUC among other non-Hispanic participants. Complete prediction performance results are provided in Supplementary Table S10.

Overall, transfer learning improved prediction performance in both cross-national and within-country race/ethnicity settings, with larger and more consistent improvements observed across the HRS race/ethnicity groups.

### Predictor Association Patterns

Tables 3 and 4 summarize the estimated odds ratios from the optimal transfer learning models across the international aging cohorts and HRS race/ethnicity groups, respectively. Complete inference results for all target cohorts are provided in Supplementary Tables S11 and S12. We further examined whether broader patterns emerged from the estimated associations across the two transfer learning settings.

**Table 3.** Estimated odds ratios for the best target–source transfer learning models across Gateway cohorts. (^∗^95% bootstrap CI excludes 1.)

| Predictor | OR (95% CI) |
|---|---|
| **CHARLS ← KLOSA + LASI + SHARE** |  |
| Age | 0.987 (0.972–0.995)<sup>*</sup> |
| Female | 1.643 (1.373–2.281)<sup>*</sup> |
| Ever Smoked | 1.023 (0.837–1.306) |
| Diabetes Treatment | 1.019 (0.891–1.257) |
| Hypertension Medication | 1.064 (0.978–1.485) |
| Medical Insurance | 0.899 (0.281–0.953)<sup>*</sup> |
| Socioeconomic Status (PPP, original units) | 0.999994 (0.999954–1.000006) |
| **HRS ← CHARLS** |  |
| Age | 0.979 (0.973–0.987)<sup>*</sup> |
| Female | 1.450 (1.272–1.641)<sup>*</sup> |
| Ever Smoked | 1.336 (1.174–1.498)<sup>*</sup> |
| Diabetes Treatment | 0.946 (0.819–1.050) |
| Hypertension Medication | 0.901 (0.713–1.094) |
| Medical Insurance | 1.902 (1.585–2.227)<sup>*</sup> |
| Socioeconomic Status (PPP, original units) | 0.999997 (0.999992–0.999999)<sup>*</sup> |
| **KLOSA ← LASI + HAALSI + SHARE** |  |
| Age | 1.053 (1.041–1.068)<sup>*</sup> |
| Female | 1.820 (1.466–2.338)<sup>*</sup> |
| Ever Smoked | 1.034 (0.745–1.601) |
| Diabetes Treatment | 1.412 (1.012–2.013)<sup>*</sup> |
| Hypertension Medication | 1.704 (1.092–2.658)<sup>*</sup> |
| Medical Insurance | 0.938 (0.888–0.952)<sup>*</sup> |
| Socioeconomic Status (PPP, original units) | 0.999981 (0.999961–0.999996)<sup>*</sup> |
| **LASI ← CHARLS + HAALSI + ELSA** |  |
| Age | 1.027 (1.018–1.042)<sup>*</sup> |
| Female | 0.958 (0.795–1.000)<sup>*</sup> |
| Ever Smoked | 1.000 (0.816–1.049) |
| Diabetes Treatment | 1.242 (1.057–1.582)<sup>*</sup> |
| Hypertension Medication | 1.000 (0.745–1.105) |
| Medical Insurance | 1.000 (0.951–1.234) |
| Socioeconomic Status (PPP, original units) | 1.000000 (0.999976–1.000076) |
| **MHAS ← CHARLS + HAALSI + SHARE + ELSA** |  |
| Age | 1.000 (0.999–1.012) |
| Female | 1.760 (1.568–2.304)<sup>*</sup> |
| Ever Smoked | 1.000 (0.960–1.193) |
| Diabetes Treatment | 1.000 (0.620–1.000)<sup>*</sup> |
| Hypertension Medication | 1.000 (0.996–1.633) |
| Medical Insurance | 1.000 (1.000–1.246)<sup>*</sup> |
| Socioeconomic Status (PPP, original units) | 1.000000 (1.000000–1.000493)<sup>*</sup> |
| **HAALSI ← KLOSA** |  |
| Age | 1.073 (1.049–1.098)<sup>*</sup> |
| Female | 1.394 (1.057–2.168)<sup>*</sup> |
| Ever Smoked | 2.569 (0.983–5.251) |
| Diabetes Treatment | 1.316 (0.549–4.722) |
| Hypertension Medication | 1.808 (1.018–4.815)<sup>*</sup> |
| Medical Insurance | 0.878 (0.488–1.278) |
| Socioeconomic Status (PPP, original units) | 1.001554 (0.999996–1.004285) |
| **SHARE ← HRS + CHARLS + KLOSA + LASI + MHAS** |  |
| Age | 1.037 (1.024–1.050)<sup>*</sup> |
| Female | 2.339 (1.890–2.925)<sup>*</sup> |
| Ever Smoked | 2.269 (1.000–5.524) |
| Diabetes Treatment | 0.696 (0.486–0.996)<sup>*</sup> |
| Hypertension Medication | 1.819 (1.442–2.249)<sup>*</sup> |
| Medical Insurance | 0.774 (0.637–0.951)<sup>*</sup> |
| Socioeconomic Status (PPP, original units) | 1.000004 (0.999998–1.000013) |
| **ELSA ← HRS + CHARLS + MHAS + HAALSI + SHARE** |  |
| Age | 1.000 (0.975–1.035) |
| Female | 1.042 (0.685–1.912) |
| Ever Smoked | 0.851 (0.423–1.216) |
| Diabetes Treatment | 0.489 (0.267–1.000)<sup>*</sup> |
| Hypertension Medication | 5.888 (1.000–153.816)<sup>*</sup> |
| Medical Insurance | 1.000 (1.000–1.000)<sup>*</sup> |
| Socioeconomic Status (PPP, original units) | 1.000000 (0.998337–1.001234) |

**Table 4.** Estimated odds ratios for the best target–source transfer learning models across HRS race/ethnicity groups. (^∗^95% bootstrap CI excludes 1.)

| Predictor | OR (95% CI) |
|---|---|
| **Black ← Hispanic + White** |  |
| Age | 0.998 (0.981–1.012) |
| Female | 1.042 (0.815–1.339) |
| Education: GED | 1.030 (0.574–1.653) |
| Education: HS Graduate | 0.709 (0.464–0.988)<sup>*</sup> |
| Education: Some College | 0.593 (0.388–0.850)<sup>*</sup> |
| Education: College+ | 0.338 (0.184–0.552)<sup>*</sup> |
| Wealth (per \$100k) | 0.963 (0.892–0.997)<sup>*</sup> |
| Smoking: Former | 0.809 (0.520–0.990)<sup>*</sup> |
| Smoking: Never | 0.695 (0.441–0.987)<sup>*</sup> |
| BMI | 1.000 (0.981–1.015) |
| Diabetes Tx: Oral only | 0.980 (0.800–1.311) |
| Diabetes Tx: Any insulin | 1.329 (1.037–2.035)<sup>*</sup> |
| Medicare | 0.979 (0.815–1.387) |
| Medicaid | 1.202 (0.960–1.646) |
| Private insurance | 0.840 (0.625–1.075) |
| **Hispanic ← White + Black** |  |
| Age | 0.994 (0.974–1.006) |
| Female | 1.208 (1.002–1.668)<sup>*</sup> |
| Education: GED | 0.745 (0.354–1.129) |
| Education: HS Graduate | 0.739 (0.490–0.991)<sup>*</sup> |
| Education: Some College | 0.390 (0.260–0.606)<sup>*</sup> |
| Education: College+ | 0.351 (0.164–0.646)<sup>*</sup> |
| Wealth (per \$100k) | 0.932 (0.842–0.981)<sup>*</sup> |
| Smoking: Former | 0.981 (0.683–1.373) |
| Smoking: Never | 0.981 (0.719–1.461) |
| BMI | 1.000 (0.976–1.015) |
| Diabetes Tx: Oral only | 0.796 (0.583–0.988)<sup>*</sup> |
| Diabetes Tx: Any insulin | 1.188 (0.890–1.801) |
| Medicare | 0.978 (0.719–1.303) |
| Medicaid | 1.228 (0.949–1.702) |
| Private insurance | 0.671 (0.468–0.923)<sup>*</sup> |
| **Other ← White + Black** |  |
| Age | 0.999 (0.999–1.046) |
| Female | 1.026 (0.580–1.234) |
| Education: GED | 1.033 (0.160–1.053) |
| Education: HS Graduate | 1.002 (0.692–1.771) |
| Education: Some College | 0.982 (0.539–1.308) |
| Education: College+ | 0.939 (0.427–0.995)<sup>*</sup> |
| Wealth (per \$100k) | 0.998 (0.982–1.018) |
| Smoking: Former | 0.992 (0.917–1.861) |
| Smoking: Never | 0.972 (0.568–1.153) |
| BMI | 1.000 (0.970–1.030) |
| Diabetes Tx: Oral only | 0.990 (0.831–1.773) |
| Diabetes Tx: Any insulin | 1.032 (0.273–1.045) |
| Medicare | 1.246 (0.993–2.824) |
| Medicaid | 1.228 (1.030–3.308)<sup>*</sup> |
| Private insurance | 0.777 (0.366–0.985)<sup>*</sup> |
| **White ← Hispanic + Black** |  |
| Age | 1.008 (0.994–1.025) |
| Female | 1.261 (1.026–1.566)<sup>*</sup> |
| Education: GED | 0.806 (0.501–1.375) |
| Education: HS Graduate | 0.691 (0.508–0.995)<sup>*</sup> |
| Education: Some College | 0.554 (0.398–0.941)<sup>*</sup> |
| Education: College+ | 0.448 (0.313–0.762)<sup>*</sup> |
| Wealth (per \$100k) | 0.991 (0.971–1.000) |
| Smoking: Former | 0.530 (0.374–0.987)<sup>*</sup> |
| Smoking: Never | 0.471 (0.333–0.877)<sup>*</sup> |
| BMI | 0.997 (0.980–1.012) |
| Diabetes Tx: Oral only | 0.960 (0.753–1.225) |
| Diabetes Tx: Any insulin | 1.225 (0.925–1.681) |
| Medicare | 0.774 (0.533–1.024) |
| Medicaid | 1.441 (1.100–2.136)<sup>*</sup> |
| Private insurance | 0.745 (0.580–0.936)<sup>*</sup> |

Predictor associations were not equally preserved across populations. Female sex showed predominantly positive associations in both the international cohorts and HRS race/ethnicity groups, although statistical significance varied across target populations. In contrast, age demonstrated consistent associations across the international cohorts but showed little evidence of association within HRS race/ethnicity groups. Similar differences in transferability were observed for other predictors, indicating that association transferability differed across predictor domains.

Greater population heterogeneity was observed for predictors related to health behaviors, disease management, and healthcare utilization. For example, smoking history showed limited and inconsistent associations across the international cohorts but was associated with the outcome in several HRS race/ethnicity groups. Similar variation was observed for other healthcare-related predictors. Notably, within HRS, health insurance was represented using several insurance categories, which exhibited relatively consistent association patterns across race/ethnicity groups. In contrast, the corresponding insurance variable in the international analysis was represented more coarsely and showed little evidence of consistent associations across cohorts.

Socioeconomic associations also differed according to the level at which populations were compared. Across the international cohorts, the country-level socioeconomic indicator (PPP) showed limited evidence of meaningful associations, whereas within HRS, individual-level socioeconomic measures demonstrated more consistent associations across race/ethnicity groups, particularly for educational attainment.

Overall, despite representing two distinct transfer learning settings, the international cohort and HRS analyses revealed remarkably similar patterns of population heterogeneity. The extent to which predictor–outcome associations were preserved varied substantially across predictors.

## Discussion

Accurate prediction of patient-reported health outcomes is increasingly important for identifying high-risk individuals and informing population health strategies among older adults with diabetes ^[3]^. However, developing prediction models that maintain satisfactory performance in heterogeneous populations remains a persistent challenge in epidemiological research ^[39]^. Prediction models developed in one population often perform less well when transported to different populations ^[7]^. In the present study, we evaluated the performance of the proposed regression-based transfer learning framework across multiple heterogeneous populations. In general, the findings suggest that regression-based transfer learning is a promising approach for improving prediction across diverse populations.

Although the magnitude of improvement varied across study settings, the proposed regression-based transfer learning framework demonstrated a generally favorable pattern in both applications. This finding is noteworthy because maintaining predictive performance across different populations remains a major challenge for clinical prediction models, which often experience performance deterioration when applied beyond the populations in which they were originally developed ^[40]^. Despite substantial heterogeneity across the international aging cohorts, the proposed framework maintained or improved predictive performance for most target populations, suggesting that it can effectively leverage information shared across related populations while accommodating population-specific differences. Negative transfer was observed only for SHARE, whereas improvements were more consistent across the RAND HRS analyses. The increasing availability of harmonized international aging cohorts has created unprecedented opportunities for cross-national analyses across diverse populations ^[22]^. These resources also provide an important foundation for applying transfer learning approaches across populations in a wide range of population health applications. The present findings suggest that the proposed framework can effectively capitalize on these harmonized resources while accommodating population-specific differences. For prediction models intended for application in large population cohorts, even modest improvements in predictive performance may have considerable public health potential. By strengthening risk stratification, these improvements may support more effective population health management and healthcare resource allocation, particularly as risk stratification tools are increasingly used to identify individuals at elevated risk and guide targeted interventions ^[41,42]^. The proposed framework also has important practical advantages. It builds on information already available from related populations and does not require additional predictors or data collection. Consequently, model adaptation can be achieved with minimal implementation burden. These characteristics make this framework a practical strategy for enhancing model performance in large-scale population health applications.

Prediction models for health outcomes among adults with diabetes have often been developed and validated within specific healthcare systems or geographic regions, with relatively limited evidence regarding their transportability across diverse populations ^[43,5]^. Consequently, relatively little is known about whether predictive information can be effectively transferred across populations with substantially different demographic characteristics and healthcare systems ^[44]^. Unlike previous studies, we applied a regression-based transfer learning framework to harmonized data from multiple international aging cohorts together with complementary analyses across racial and ethnic groups within the RAND HRS. By systematically evaluating transfer learning across these complementary population settings under a unified analytical framework, this work extends its application to cross-national population health prediction.

The contrasting patterns across the two applications suggest that the effectiveness of transfer learning may depend on the degree of similarity between populations. Improvements were more consistent across racial and ethnic groups within the RAND HRS, whose participants were drawn from the same national study and shared a common survey design, measurement framework, and healthcare context ^[23]^. In contrast, the Gateway cohorts represent distinct national populations with substantial differences in health, social environments, and population characteristics, despite the use of harmonized variables to facilitate cross-national comparisons ^[22]^. The negative transfer observed for SHARE may further illustrate this challenge. As a multinational survey, SHARE encompasses populations from diverse national and institutional contexts, which may complicate information transfer ^[27]^. More broadly, these findings are consistent with previous evidence that transfer learning is less effective when the source and target domains are insufficiently related ^[21,45]^. Careful assessment of source-target compatibility may therefore be important when selecting source populations for transfer learning in heterogeneous population settings.

Our findings should be interpreted in light of several limitations. Although the present study included multiple international aging cohorts, these cohorts were not intended to represent all populations or healthcare systems. Therefore, additional validation in broader geographic regions and healthcare settings will be important to further establish the transportability of the proposed framework ^[8,46]^. We also did not explicitly identify or quantify the population characteristics that influenced transfer learning performance. A better understanding of these characteristics may inform source population selection and further enhance model transportability ^[40]^. In addition, many predictors and outcomes were derived from self-reported survey data. Although harmonized cohort studies provide unique opportunities for international comparisons, self-reported measures may provide less detailed clinical information than electronic health records ^[47]^ and may be subject to reporting error or outcome misclassification ^[48]^. Future studies integrating harmonized cohort data with richer clinical information may further improve transfer learning performance.

Despite these limitations, the transfer learning framework used in this study improved prediction across most target populations without requiring additional predictors or data collection. These findings indicate that predictive information can be effectively transferred across both international cohorts and population subgroups within a national study, suggesting that this framework is not limited to a single data structure or population setting. The same principle may be particularly valuable in hospitals or healthcare systems where local data are insufficient to support robust model development but larger, related data sources are available. Previous studies have similarly shown that transfer learning can improve prediction for smaller population groups and facilitate model adaptation across clinical sites ^[49,50]^. Further evaluation using electronic health records and other clinical data sources will help determine the broader applicability of this framework in epidemiologic and healthcare research.

## Data Availability

The data used in this study are publicly available through the Gateway to Global Aging Data and the respective cohort data repositories, including the RAND Health and Retirement Study (RAND HRS). All datasets are subject to their respective data access and use agreements. Detailed data access links are provided in the manuscript. The analytic code used in this study is publicly available on GitHub.

https://www.mhasweb.org/DataProducts/Home.aspx

https://www.iipsindia.ac.in/content/LASI-data

https://charls.charlsdata.com/pages/data/111/en.html

https://beta.ukdataservice.ac.uk/

https://survey.keis.or.kr/eng/klosa/klosa01.jsp

https://dataverse.harvard.edu/dataset.xhtml?persistentId=doi:10.7910/DVN/TW84UI

https://share-eric.eu/data/become-a-user

https://hrsdata.isr.umich.edu/data-products/rand

https://github.com/Xuzhi-Wang/Diabetes-Transfer-Learning

## Data availability

- Mexican Health and Aging Study (MHAS): https://www.mhasweb.org/DataProducts/Home.aspx.
- Longitudinal Ageing Study in India (LASI): https://www.iipsindia.ac.in/content/LASI-data.
- China Health and Retirement Longitudinal Study (CHARLS): https://charls.charlsdata.com/pages/data/111/en.html.
- English Longitudinal Study of Ageing (ELSA): https://beta.ukdataservice.ac.uk/.
- Korean Longitudinal Study of Ageing (KLoSA): https://survey.keis.or.kr/eng/klosa/klosa01.jsp.
- Health and Ageing in Africa: A Longitudinal Study of an INDEPTH Community (HAALSI): https://dataverse.harvard.edu/dataset.xhtml?persistentId=doi:10.7910/DVN/TW84UI.
- Survey of Health, Ageing and Retirement in Europe (SHARE): https://share-eric.eu/data/become-a-user.
- RAND Health and Retirement Study (RAND HRS): https://hrsdata.isr.umich.edu/data-products/rand.

All datasets are subject to their respective data access and use agreements. The analytic code used in this study is publicly available at https://github.com/Xuzhi-Wang/Diabetes-Transfer-Learning.

## Author contributions statement

XW was responsible for data preparation, coding, statistical analysis, and manuscript writing. YC and WS also contributed to data preparation. JB provided clinical and public health expertise and contributed to the interpretation of the findings. TL provided statistical and methodological guidance and supervised the project.

## Acknowledgments

We acknowledge the principal investigators, study participants, and research teams of the Health and Retirement Study (HRS), the Mexican Health and Aging Study (MHAS), the Longitudinal Ageing Study in India (LASI), the China Health and Retirement Longitudinal Study (CHARLS), the English Longitudinal Study of Ageing (ELSA), the Korean Longitudinal Study of Ageing (KLoSA), the Health and Ageing in Africa: A Longitudinal Study of an INDEPTH Community in South Africa (HAALSI), and the Survey of Health, Ageing and Retirement in Europe (SHARE) for establishing and maintaining these valuable longitudinal resources. We also acknowledge the Gateway to Global Aging Data team for supporting the harmonization and cross-cohort accessibility of these datasets, as well as the RAND HRS team for developing and maintaining the RAND HRS data product.

This work was supported in part through the NYU IT High Performance Computing resources, services, and staff expertise.

## References

1. Genitsaridi I, Salpea P, Salim A, et al. 11th edition of the IDF Diabetes Atlas: global, regional, and national diabetes prevalence estimates for 2024 and projections for 2050. The Lancet Diabetes & Endocrinology, 14(2):149–156, February 2026.

2. American Diabetes Association Professional Practice Committee for Diabetes*, Bajaj M, McCoy RG, et al. 13. Older Adults: Standards of Care in Diabetes—2026. Diabetes Care, 49(Supplement 1):S277–S296, January 2026.

3. Terwee CB, Elders PJM, Blom MT, et al. Patient-reported outcomes for people with diabetes: what and how to measure? A narrative review. Diabetologia, 66(8):1357–1377, August 2023.

4. Slieker RC, Van Der Heijden AAWA, Siddiqui MK, et al. Performance of prediction models for nephropathy in people with type 2 diabetes: systematic review and external validation study. BMJ, page 2134, September 2021.

5. Ghafoor B, Zaccardi F, Khunti K, et al. Comparative predictive accuracy of machine learning versus traditional statistical methods for diabetes-related complications: A systematic review and analysis. Diabetic Medicine, page e70398, June 2026.

6. De Hond AAH, Shah VB, Kant IMJ, et al. Perspectives on validation of clinical predictive algorithms. npj Digital Medicine, 6(1):86, May 2023.

7. Collins GS, Dhiman P, Ma J, et al. Evaluation of clinical prediction models (part 1): from development to external validation. BMJ, 384:e074819, January 2024.

8. Riley RD, Archer L, Snell KIE, et al. Evaluation of clinical prediction models (part 2): how to undertake an external validation study. BMJ, 384:e074820, January 2024.

9. Yang J, Soltan AAS, Clifton DA. Machine learning generalizability across healthcare settings: insights from multi-site COVID-19 screening. npj Digital Medicine, 5(1):69, June 2022.

10. Lasko TA, Strobl EV, Stead WW. Why do probabilistic clinical models fail to transport between sites. npj Digital Medicine, 7(1):53, March 2024.

11. Yordanov TR, Lopes RR, Ravelli AC, et al. An integrated approach to geographic validation helped scrutinize prediction model performance and its variability. Journal of Clinical Epidemiology, 157:13–21, May 2023.

12. De Jong VMT, Hoogland J, Moons KGM, et al. Propensity-based standardization to enhance the validation and interpretation of prediction model discrimination for a target population. Statistics in Medicine, 42(19):3508–3528, August 2023.

13. Li M, Tan A, Song L, et al. Performance of diabetes risk prediction models: a systematic review and meta-analysis. Endocrine Connections, 14(11):e250353, November 2025.

14. Tan KR, Seng JJB, Kwan YH, et al. Evaluation of Machine Learning Methods Developed for Prediction of Diabetes Complications: A Systematic Review. Journal of Diabetes Science and Technology, 17(2):474–489, March 2023.

15. Riley RD, Ensor J, Snell KIE, et al. Calculating the sample size required for developing a clinical prediction model. BMJ, page m441, March 2020.

16. Efthimiou O, Seo M, Chalkou K, et al. Developing clinical prediction models: a step-by-step guide. BMJ, 386:e078276, September 2024.

17. Li S, Cai T, Duan R. Targeting underrepresented populations in precision medicine: A federated transfer learning approach. The Annals of Applied Statistics, 17(4), December 2023.

18. Tian Y, Feng Y. Transfer Learning Under High-Dimensional Generalized Linear Models. Journal of the American Statistical Association, 118(544):2684–2697, October 2023.

19. Gu T, Lee PH, Duan R. COMMUTE: Communication-efficient transfer learning for multi-site risk prediction. Journal of Biomedical Informatics, 137:104243, January 2023.

20. Li S, Li X, Yu K, et al. Bridging Data Gaps in Healthcare: A Scoping Review of Transfer Learning in Structured Data Analysis. Health Data Science, 5:0321, January 2025.

21. Gu T, Han Y, Duan R. Robust angle-based transfer learning in high dimensions. Journal of the Royal Statistical Society Series B: Statistical Methodology, 87(3):723–745, July 2025.

22. Lee J, Phillips D, Wilkens J, et al. Gateway to Global Aging Data: Resources for Cross-National Comparisons of Family, Social Environment, and Healthy Aging. The Journals of Gerontology: Series B, 76(Supplement 1):S5–S16, June 2021.

23. Sonnega A, Faul JD, Ofstedal MB, et al. Cohort Profile: the Health and Retirement Study (HRS). International Journal of Epidemiology, 43(2):576–585, April 2014.

24. Wong R, Michaels-Obregon A, Palloni A. Cohort Profile: The Mexican Health and Aging Study (MHAS). International Journal of Epidemiology, 46(2):e2–e2, April 2017.

25. Perianayagam A, Bloom D, Lee J, et al. Cohort Profile: The Longitudinal Ageing Study in India (LASI). International Journal of Epidemiology, 51(4):e167–e176, August 2022.

26. Zhao Y, Hu Y, Smith JP, et al. Cohort Profile: The China Health and Retirement Longitudinal Study (CHARLS). International Journal of Epidemiology, 43(1):61–68, February 2014.

27. Böorsch-Supan A, Brandt M, Hunkler C, et al. Data Resource Profile: The Survey of Health, Ageing and Retirement in Europe (SHARE). International Journal of Epidemiology, 42(4):992–1001, August 2013.

28. Steptoe A, Breeze E, Banks J, et al. Cohort Profile: The English Longitudinal Study of Ageing. International Journal of Epidemiology, 42(6):1640–1648, December 2013.

29. Jang SN. Korean Longitudinal Study of Ageing (KLoSA): Overview of Research Design and Contents. In Pachana NA, editor, Encyclopedia of Geropsychology, pages 1–9. Springer Singapore, Singapore, 2016.

30. Góomez-Olivé FX, Montana L, Wagner RG, et al. Cohort Profile: Health and Ageing in Africa: A Longitudinal Study of an INDEPTH Community in South Africa (HAALSI). International Journal of Epidemiology, 47(3):689–690j, June 2018.

31. Health and Retirement Study. RAND HRS Longitudinal File 2018 (V2), July 2022.

32. RAND Center for the Study of Aging. RAND HRS Longitudinal File 2018 (V2), July 2022.

33. World Bank. Purchasing Power Parities and the Size of World Economies: Results from the 2021 International Comparison Program. World Bank, 2024.

34. Buuren SV, Groothuis-Oudshoorn K. mice : Multivariate Imputation by Chained Equations in R. Journal of Statistical Software, 45(3), 2011.

35. Lee J, Phillips D, Wilkens J, et al. Cross-Country Comparisons of Disability and Morbidity: Evidence from the Gateway to Global Aging Data. The Journals of Gerontology: Series A, 73(11):1519–1524, October 2018.

36. Verbeek E, Drewes Y, Gussekloo J. Visual impairment as a predictor for deterioration in functioning: the Leiden 85-plus Study. BMC Geriatrics, 22(1):397, December 2022.

37. Tian Y, Feng Y. glmtrans: Transfer Learning under Regularized Generalized Linear Models, April 2021. Institution: Comprehensive R Archive Network Pages: 2.1.0.

38. Friedman J, Hastie T, Tibshirani R. Regularization Paths for Generalized Linear Models via Coordinate Descent. Journal of Statistical Software, 33(1), 2010.

39. De Jong VMT, Moons KGM, Eijkemans MJC, et al. Developing more generalizable prediction models from pooled studies and large clustered data sets. Statistics in Medicine, 40(15):3533–3559, July 2021.

40. Steingrimsson JA, Gatsonis C, Li B, et al. Transporting a Prediction Model for Use in a New Target Population. American Journal of Epidemiology, 192(2):296–304, February 2023.

41. Ravaut M, Sadeghi H, Leung KK, et al. Predicting adverse outcomes due to diabetes complications with machine learning using administrative health data. npj Digital Medicine, 4(1):24, February 2021.

42. Oddy C, Zhang J, Morley J, et al. Promising algorithms to perilous applications: a systematic review of risk stratification tools for predicting healthcare utilisation. BMJ Health & Care Informatics, 31(1):e101065, June 2024.

43. Ndjaboue R, Ngueta G, Rochefort-Brihay C, et al. Prediction models of diabetes complications: a scoping review. Journal of Epidemiology and Community Health, 76(10):896–904, October 2022.

44. Kirchler M, Ferro M, Lorenzini V, et al. Large language models improve transferability of electronic health record-based predictions across countries and coding systems. npj Digital Medicine, 9(1):177, January 2026.

45. Peng Y, Wang L. Heterogeneity-aware transfer learning for high-dimensional linear regression models. Computational Statistics & Data Analysis, 206:108129, June 2025.

46. Pfeiffer RM, Chen Y, Gail MH, et al. Accommodating population differences when validating risk prediction models. Statistics in Medicine, 41(24):4756–4780, October 2022.

47. Ghildayal N, Nagavedu K, Wiltz JL, et al. Public Health Surveillance in Electronic Health Records: Lessons From PCORnet. Preventing Chronic Disease, 21:230417, July 2024.

48. Potter T, Almidani L, Diaz M, et al. Concordance between Self-Reported Visual Difficulty and Objective Visual Impairment. Ophthalmology, 131(12):1447–1456, December 2024.

49. Cao J, Balczewski E, Ladhania R, et al. Exploring the limits of localization: federated model stacking improves hospital-level prediction in a national research network. npj Digital Medicine, 9(1):492, April 2026.

50. Kim JH, Glymour MM, Langa KM, et al. Improving accuracy in the estimation of probable dementia in racially and ethnically diverse groups with penalized regression and transfer learning. American Journal of Epidemiology, 195(1):237–245, January 2026.

