## Supplementary Material for "Improving Prediction of Patient-Reported Health Outcomes Among Older Adults With Diabetes Across Diverse Populations Through Transfer Learning"

Table S1: Distribution of diabetes status by cohort before participant selection.

| Cohort | Sample Size | Diagnosed Diabetes | No Diagnosed Diabetes | Missing |
|---|---|---|---|---|
| HRS | 15 064 | 4449 (29.53%) | 10 613 (70.45%) | 2 (0.02%) |
| MHAS | 12 931 | 3488 (26.97%) | 9421 (72.86%) | 22 (0.17%) |
| LASI | 42 069 | 6301 (14.98%) | 35 660 (84.77%) | 108 (0.25%) |
| CHARLS | 14 099 | 2 127 (15.09%) | 11 322 (80.30%) | 650 (4.61%) |
| ELSA | 7349 | 269 (3.66%) | 7080 (96.34%) | 0 (0.00%) |
| KLoSA | 6940 | 1382 (19.91%) | 5558 (80.09%) | 0 (0.00%) |
| HAALSI | 5059 | 581 (11.48%) | 2695 (53.27%) | 1783 (35.24%) |
| SHARE | 72 088 | 10 367 (14.38%) | 61 233 (84.94%) | 488 (0.68%) |

Diabetes status was determined using self-reported physician diagnosis variables available in each survey prior to data harmonization, imputation, and the application of eligibility criteria. Percentages were calculated using the total number of respondents within each original dataset. Missing indicates participants with unavailable responses for the diabetes diagnosis variable in the original datasets. The relatively high proportion of missing diabetes diagnosis data observed in HAALSI reflects unavailable responses in the original survey and was addressed during subsequent eligibility assessment and data processing.

Table S2: Overview of harmonized variables included in the Gateway analysis.

| Domain | Variables | Measurement type |
|---|---|---|
| Demographic characteristics | Age; Female sex | Continuous; Binary |
| Socioeconomic characteristics | PPP-adjusted socioeconomic indicator; Public insurance | Continuous; Binary |
| Behavioural factors | Ever smoking | Binary |
| Diabetes management | Diabetes treatment | Binary |
| Cardiometabolic management | Antihypertensive medication | Binary |
| Health-related outcome | Pain and/or visual impairment | Binary (0 = neither; 1 = either or both) |

The socioeconomic indicator was converted to international dollars using country-specific 2018 Purchasing Power Parity (PPP) conversion factors.

Table S3: Missingness of selected variables in SHARE before outcome exclusion and predictor imputation.

| Variable | Missing, n | Missing, % | Available, n | Available, % |
|---|---|---|---|---|
| Age | 1 | 0.01 | 10 366 | 99.99 |
| Gender | 0 | 0.00 | 10 367 | 100.00 |
| Ever smoke | 10 286 | 99.22 | 81 | 0.78 |
| Public medical insurance | 8535 | 82.33 | 1832 | 17.67 |
| Diabetes treatment | 23 | 0.22 | 10 344 | 99.78 |
| Hypertension medication | 23 | 0.22 | 10 344 | 99.78 |
| Pain | 8242 | 79.50 | 2125 | 20.50 |
| Visual impairment | 8243 | 79.51 | 2124 | 20.49 |
| Pension income | 0 | 0.00 | 10 367 | 100.00 |
| Social welfare | 0 | 0.00 | 10 367 | 100.00 |

Missingness was assessed among participants with diagnosed diabetes before exclusion for missing outcome data and before predictor imputation. Pain and visual impairment were not imputed; participants missing either outcome component were excluded before construction of the binary outcome.

Table S4: Detailed distribution of categorical study variables among participants with diagnosed diabetes across the eight cohorts.

| Variable | Category | HRS | MHAS | LASI | CHARLS | ELSA | KLoSA | HAALSI | SHARE |
|---|---|---|---|---|---|---|---|---|---|
| Ever Smoking | Yes, n (%) | 2498 (56.15) | 1245 (35.69) | 1681 (26.68) | 841 (39.54) | 165 (61.3) | 107(7.74) | 52 (10.2) | 29(0.28) |
| | No, n (%) | 1950 (43.83) | 2240 (64.22) | 4569 (72.51) | 1283 (60.32) | 90 (33.5) | 1275(92.26) | 455 (89.4) | 52(0.50) |
| | Missing, n (%) | 1(0.02) | 3 (0.09) | 51 (0.81) | 3 (0.14) | 14 (5.2) | 0(0) | 2 (0.4) | 10 286(99.22) |
| Diabetes Treatment | Yes, n (%) | 2992(67.25) | 3242(92.95) | 5253(83.37) | 675(31.73) | 127 (47.2) | 1154(83.5) | 162 (31.8) | 9150(88.26) |
| | No, n (%) | 1193(26.82) | 246(7.05) | 1047(16.62) | 441(20.73) | 96 (35.7) | 228(16.5) | 13 (2.6) | 1194(11.52) |
| | Missing, n (%) | 264(5.93) | 0(0) | 1(0.02) | 1011(47.53) | 46 (17.1) | 0(0) | 334 (65.6) | 23(0.22) |
| Antihypertensive Med | Yes, n (%) | 3321(74.65) | 2162(61.98) | 3569(56.64) | 1082(50.87) | 128 (47.6) | 1246(90.16) | 261 (51.3) | 7625(73.55) |
| | No, n (%) | 273(6.14) | 152(4.36) | 543(8.62) | 267(12.55) | 4 (1.5) | 136(9.84) | 24 (4.7) | 2,719(26.23) |
| | Missing, n (%) | 855(19.22) | 1174(33.66) | 2189(34.74) | 778(36.58) | 137 (50.9) | 0(0) | 224 (44.0) | 23(0.22) |
| Pain | Yes, n (%) | 2136(48.01) | 1474(42.26) | 3315(52.61) | 1442(67.80) | 115 (42.8) | 577(41.75) | 161 (31.6) | 1,248(12.04) |
| | No, n (%) | 2297(51.63) | 1697(48.65) | 2956(46.91) | 682(32.06) | 99 (36.8) | 805(58.25) | 313 (61.5) | 877(8.46) |
| | Missing, n (%) | 16(0.36) | 317(9.09) | 30(0.48) | 3(0.14) | 55 (20.4) | 0(0) | 35 (6.9) | 8,242(79.50) |
| Visual Impairment | Yes, n (%) | 577(12.97) | 351(10.06) | 1577(25.03) | 702(33.00) | 6 (2.2) | 90(6.51) | 164 (32.2) | 240(2.32) |
| | No, n (%) | 3869(86.96) | 2811(80.59) | 4717(74.86) | 1422(66.85) | 263 (97.8) | 1282(92.76) | 344 (67.6) | 1884(18.17) |
| | Missing, n (%) | 3(0.07) | 326(9.35) | 7(0.11) | 3(0.14%) | 0(0) | 10(0.72) | 1 (0.2) | 8243(79.51) |
| Public Insurance | Yes, n (%) | 3326(74.76) | 1857(53.24) | 1386(22.00) | 2078(97.7) | NA | 1382(100) | 62 (12.2) | 1027(9.91) |
| | No, n (%) | 1084(24.37) | 1623(46.53) | 4858(77.10) | 46(2.16) | NA | 0(0) | 446 (87.6) | 805(7.77) |
| | Missing, n (%) | 39(0.88%) | 8(0.23) | 57(0.90%) | 3(0.14) | NA | 0(0) | 1 (0.2) | 8535(82.33) |
| Female Sex | Yes, n (%) | 2535(56.98) | 2140(61.35) | 2084(33.07) | 1176(55.29) | 128 (47.6) | 774(56.01) | 299 (58.7) | 5438(52.45) |
| | No, n (%) | 1914(43.02) | 1348(38.65) | 4217(66.93) | 951(44.71) | 141 (52.4) | 608(43.99) | 210 (41.3) | 4929(47.55) |
| | Missing, n (%) | 0(0) | 0(0) | 0(0) | 0(0) | 0(0) | 0(0) | 0(0) | 0(0) |

Values are presented as number of participants and percentages within each cohort. Percentages were calculated using the total number of eligible participants with diagnosed diabetes in each cohort. Missing indicates unavailable responses in the harmonized datasets before predictor imputation. Public health insurance coverage was not directly assessed in ELSA because the United Kingdom provides universal healthcare coverage; therefore, values are reported as NA. Several variables exhibited substantial missingness in specific cohorts because these measures were not collected in all survey waves or were unavailable within the harmonized datasets. Missing predictor values were subsequently addressed according to the cohort–specific imputation strategies described in the Methods. Pain and visual impairment were not imputed.

Table S5: Detailed distribution of continuous study variables among participants with diagnosed diabetes across the eight cohorts.

| Variable | Summary | HRS | MHAS | LASI | CHARLS | ELSA | KLoSA | HAALSI | SHARE |
|---|---|---|---|---|---|---|---|---|---|
| Pension Income | Mean (SD) | 23,683.39 (82,666.81) | 378.98 (595.51) | 546.18 (1505.97) | 3548.47 (4595.11) | 77.76 (115.77) | 336.35 (552.06) | 1774.37 (823.20) | 2342.404 (22132.75) |
| | Median (IQR) | 13,572 [5022, 28,629] | 215.60 [118.58, 404.74] | 0 [0, 195.36] | 488.51 [244.25, 6350.62] | 27.71 [0, 122.17] | 241 [0, 373] | 1700 [1700, 1700] | 0 [0, 0] |
| | Missing, n (%) | 3611 (81.16) | 2477 (71.01) | 216 (3.43) | 825 (38.79) | 12 (4.46) | 0 (0) | 115 (22.59) | 0 (0) |
| Social Welfare | Mean (SD) | 14,293.04 (11 681.49) | 62.80 (73.25) | 72.21 (269.43) | 879.04 (3223.23) | 18.97 (38.99) | 688.83 (183.34) | 1022.4 (714.1) | 66.83 (1553.45) |
| | Median (IQR) | 13,200 [9000, 18,000] | 53.9 [26.65, 62.52] | 7.33 [0, 46.89] | 256.47 [129.45, 586.21] | 0 [0, 17.5] | 673 [553, 841] | 800 [410, 1230] | 0 [0, 0] |
| | Missing, n (%) | 1688 (37.94) | 2333 (66.89) | 216 (3.43) | 1936 (91.02) | 12 (4.46) | 1335 (96.60) | 365 (71.7) | 0 (0) |
| Age | Mean (SD) | 69.91 (9.82) | 68.26 (8.66) | 66.03 (7.72) | 66.55 (8.19) | 69.91 (9.26) | 73.12 (8.99) | 70.3 (10.0) | 71.79 (8.67) |
| | Median (IQR) | 68 [61, 77] | 68 [61, 74] | 65 [60, 71] | 65 [60, 72] | 70 [63, 77] | 73 [66, 80] | 69 [62, 76] | 71 [65, 78] |
| | Missing, n (%) | 0 (0) | 0 (0) | 0 (0) | 0 (0) | 0 (0) | 0 (0) | 0 (0) | 1 (0.0096) |

Continuous variables are summarized as mean (standard deviation) and median (interquartile range). Percentages of missing values were calculated within each cohort before predictor imputation. Pension income and social welfare benefits are reported as 2018 Purchasing Power Parity (PPP)-adjusted U.S. dollars using country-specific PPP conversion factors to facilitate cross-cohort comparisons. For cohorts in which these measures were available only at the household level, household-level amounts were divided by household size to derive per-capita estimates.

Table S6: Missingness of the outcome, race/ethnicity, and predictors in the RAND Health and Retirement Study before complete-case analysis.

| Category | Variable | Missing, n | Missing, % | Available, n | Available, % |
|---|---|---|---|---|---|
| Outcome | Self-reported eyesight | 11 | 0.26 | 4,248 | 99.74 |
| Domain variable | Race/ethnicity | 5 | 0.12 | 4,254 | 99.88 |
| Predictors | Age | 0 | 0.00 | 4,259 | 100.00 |
| | Female sex | 0 | 0.00 | 4,259 | 100.00 |
| | Education | 0 | 0.00 | 4,259 | 100.00 |
| | Wealth | 0 | 0.00 | 4,259 | 100.00 |
| | Smoking status | 16 | 0.38 | 4,243 | 99.62 |
| | BMI | 67 | 1.57 | 4,192 | 98.43 |
| | Diabetes treatment | 121 | 2.84 | 4,138 | 97.16 |
| | Medicare | 45 | 1.06 | 4,214 | 98.94 |
| | Medicaid | 80 | 1.88 | 4,179 | 98.12 |
| | Private insurance | 89 | 2.09 | 4,170 | 97.91 |

Missingness was assessed among the 4,259 participants with diagnosed diabetes before complete-case exclusion. Participants with missing outcome, race/ethnicity, or predictor data were excluded, resulting in a final analytic sample of 3,926 participants. Missingness across variables was not mutually exclusive.

Table S7: Transferable source cohorts identified for each target cohort in the Gateway analysis.

| Target cohort | Source cohort | Source loss | Transferability threshold | Loss difference |
|---|---|---|---|---|
| CHARLS | SHARE | 0.582 | 0.604 | -0.022 |
| | ELSA | 0.584 | 0.604 | -0.020 |
| | HAALSI | 0.586 | 0.604 | -0.018 |
| | HRS | 0.595 | 0.604 | -0.009 |
| HRS | ELSA | 0.680 | 0.699 | -0.020 |
| | HAALSI | 0.683 | 0.699 | -0.016 |
| | KLoSA | 0.684 | 0.699 | -0.016 |
| | CHARLS | 0.685 | 0.699 | -0.015 |
| | MHAS | 0.685 | 0.699 | -0.014 |
| | SHARE | 0.690 | 0.699 | -0.009 |
| KLoSA | HAALSI | 0.651 | 0.684 | -0.033 |
| | ELSA | 0.662 | 0.684 | -0.022 |
| | SHARE | 0.673 | 0.684 | -0.011 |
| | MHAS | 0.679 | 0.684 | -0.005 |
| | HRS | 0.684 | 0.684 | 0.000 |
| LASI | ELSA | 0.414 | 0.434 | -0.019 |
| | CHARLS | 0.418 | 0.434 | -0.015 |
| | HAALSI | 0.419 | 0.434 | -0.015 |
| | SHARE | 0.424 | 0.434 | -0.010 |
| MHAS | ELSA | 0.683 | 0.704 | -0.021 |
| | KLoSA | 0.683 | 0.704 | -0.020 |
| | HRS | 0.684 | 0.704 | -0.020 |
| | HAALSI | 0.686 | 0.704 | -0.018 |
| | SHARE | 0.687 | 0.704 | -0.016 |
| | CHARLS | 0.694 | 0.704 | -0.010 |
| HAALSI | KLoSA | 0.648 | 0.667 | -0.019 |
| | ELSA | 0.663 | 0.667 | -0.004 |
| SHARE | ELSA | 0.631 | 0.649 | -0.019 |
| | HAALSI | 0.632 | 0.649 | -0.017 |
| | KLoSA | 0.638 | 0.649 | -0.012 |
| | CHARLS | 0.645 | 0.649 | -0.004 |
| ELSA | MHAS | 0.688 | 0.709 | -0.021 |
| | HRS | 0.691 | 0.709 | -0.018 |
| | SHARE | 0.696 | 0.709 | -0.013 |
| | HAALSI | 0.700 | 0.709 | -0.009 |

Table S8: Transferable source cohorts identified for each target racial/ethnic group in the HRS cohort.

| Target cohort | Source cohort | Source loss | Transferability threshold | Loss difference |
|---|---|---|---|---|
| Black | Hispanic | 0.637 | 0.666 | -0.029 |
| | Other | 0.647 | 0.666 | -0.019 |
| | White | 0.662 | 0.666 | -0.003 |
| Hispanic | Black | 0.664 | 0.689 | -0.025 |
| | Other | 0.667 | 0.689 | -0.022 |
| Other | Black | 0.607 | 0.633 | -0.026 |
| | White | 0.625 | 0.633 | -0.007 |
| White | Black | 0.502 | 0.527 | -0.025 |
| | Hispanic | 0.507 | 0.527 | -0.020 |
| | Other | 0.508 | 0.527 | -0.020 |

Table S9: Prediction performance of the optimal transferable source combinations for each target cohort in the Gateway cohorts.

| Target cohort | Source combination | No. of source cohorts | AUC | | | Brier score | | | ΔAUC (TL vs Baseline) | | | ΔAUC (TL vs Pooled) | | |
|---|---|---|---|---|---|---|---|---|---|---|---|---|---|---|
| | | | Baseline | Pooled | Transfer | Baseline | Pooled | Transfer | Mean | Lower 95% CI | Upper 95% CI | Mean | Lower 95% CI | Upper 95% CI |
| CHARLS | KLoSA+LASI+SHARE | 3 | 0.595 | 0.540 | 0.600 | 0.194 | 0.199 | 0.194 | 0.006 | 0.005 | 0.006 | 0.061 | 0.058 | 0.064 |
| HRS | CHARLS | 1 | 0.599 | 0.596 | 0.601 | 0.242 | 0.242 | 0.242 | 0.002 | 0.002 | 0.002 | 0.004 | 0.004 | 0.005 |
| KLoSA | LASI+HAALSI+SHARE | 3 | 0.673 | 0.674 | 0.674 | 0.226 | 0.234 | 0.226 | 0.001 | 0.000 | 0.001 | 0.000 | -0.001 | 0.001 |
| LASI | CHARLS+HAALSI+ELSA | 3 | 0.572 | 0.543 | 0.573 | 0.123 | 0.123 | 0.123 | 0.001 | 0.001 | 0.002 | 0.030 | 0.028 | 0.032 |
| MHAS | CHARLS+HAALSI+SHARE+ELSA | 4 | 0.587 | 0.586 | 0.592 | 0.244 | 0.244 | 0.244 | 0.005 | 0.004 | 0.006 | 0.006 | 0.005 | 0.007 |
| HAALSI | KLoSA | 1 | 0.699 | 0.697 | 0.703 | 0.222 | 0.223 | 0.221 | 0.003 | 0.002 | 0.005 | 0.006 | 0.004 | 0.008 |
| SHARE | HRS+CHARLS+KLoSA+LASI+MHAS | 5 | 0.665 | 0.625 | 0.665 | 0.216 | 0.224 | 0.216 | 0.000 | 0.000 | 0.000 | 0.040 | 0.038 | 0.042 |
| ELSA | HRS+CHARLS+MHAS+HAALSI+SHARE | 5 | 0.580 | 0.566 | 0.585 | 0.244 | 0.245 | 0.244 | 0.005 | 0.000 | 0.010 | 0.019 | 0.011 | 0.027 |



Table S10: Prediction performance of the optimal transferable source combinations for each target cohort in the HRS racial/ethnic subgroups.

| Target cohort | Source combination | No. of source cohorts | AUC | | | Brier score | | | ΔAUC (TL vs Baseline) | | | ΔAUC (TL vs Pooled) | | |
|---|---|---|---|---|---|---|---|---|---|---|---|---|---|---|
| | | | Baseline | Pooled | Transfer | Baseline | Pooled | Transfer | Mean | Lower 95% CI | Upper 95% CI | Mean | Lower 95% CI | Upper 95% CI |
| Black | Hispanic+White | 2 | 0.629 | 0.649 | 0.639 | 0.223 | 0.219 | 0.221 | 0.011 | 0.009 | 0.012 | -0.009 | -0.010 | -0.008 |
| Hispanic | White+Black | 2 | 0.652 | 0.670 | 0.662 | 0.232 | 0.229 | 0.231 | 0.010 | 0.008 | 0.011 | -0.008 | -0.010 | -0.007 |
| Other | White+Black | 2 | 0.564 | 0.590 | 0.595 | 0.213 | 0.213 | 0.208 | 0.031 | 0.023 | 0.038 | 0.005 | 0.000 | 0.010 |
| White | Hispanic+Black | 2 | 0.620 | 0.638 | 0.628 | 0.158 | 0.156 | 0.158 | 0.008 | 0.006 | 0.009 | -0.010 | -0.011 | -0.010 |

Table S11: Inference results for the optimal transferable models in the Gateway cohorts.

| Target cohort | Source combination | Predictor | OR | 95% CI | Significant |
|---|---|---|---|---|---|
| CHARLS | KLoSA+LASI+SHARE | Age | 0.987 | 0.972-0.995 | Yes |
| | | Female | 1.643 | 1.373-2.281 | Yes |
| | | Ever Smoked | 1.023 | 0.837-1.306 | No |
| | | Diabetes Treatment | 1.019 | 0.891-1.257 | No |
| | | Hypertension Medication | 1.064 | 0.978-1.485 | No |
| | | Medical Insurance | 0.899 | 0.281-0.953 | Yes |
| | | Socioeconomic status (PPP) | 0.999994 | 0.999954-1.000006 | No |
| HRS | CHARLS | Age | 0.979 | 0.973-0.987 | Yes |
| | | Female | 1.450 | 1.272-1.641 | Yes |
| | | Ever Smoked | 1.336 | 1.174-1.498 | Yes |
| | | Diabetes Treatment | 0.946 | 0.819-1.050 | No |
| | | Hypertension Medication | 0.901 | 0.713-1.094 | No |
| | | Medical Insurance | 1.902 | 1.585-2.227 | Yes |
| | | Socioeconomic status (PPP) | 0.999997 | 0.999992-0.999999 | Yes |
| KLoSA | LASI+HAALSI+SHARE | Age | 1.053 | 1.041-1.068 | Yes |
| | | Female | 1.820 | 1.466-2.338 | Yes |
| | | Ever Smoked | 1.034 | 0.745-1.601 | No |
| | | Diabetes Treatment | 1.412 | 1.012-2.013 | Yes |
| | | Hypertension Medication | 1.704 | 1.092-2.658 | Yes |
| | | Medical Insurance | 0.938 | 0.888-0.952 | Yes |
| | | Socioeconomic status (PPP) | 0.999981 | 0.999961-0.999996 | Yes |
| LASI | CHARLS+HAALSI+ELSA | Age | 1.027 | 1.018-1.042 | Yes |
| | | Female | 0.958 | 0.795-1.000 | Yes |
| | | Ever Smoked | 1.000 | 0.816-1.049 | No |
| | | Diabetes Treatment | 1.242 | 1.057-1.582 | Yes |
| | | Hypertension Medication | 1.000 | 0.745-1.105 | No |
| | | Medical Insurance | 1.000 | 0.951-1.234 | No |
| | | Socioeconomic status (PPP) | 1.000000 | 0.999976-1.000076 | No |
| MHAS | CHARLS+HAALSI+ SHARE+ELSA | Age | 1.000 | 0.999-1.012 | No |
| | | Female | 1.760 | 1.568-2.304 | Yes |
| | | Ever Smoked | 1.000 | 0.960-1.193 | No |
| | | Diabetes Treatment | 1.000 | 0.620-1.000 | Yes |
| | | Hypertension Medication | 1.000 | 0.996-1.633 | No |
| | | Medical Insurance | 1.000 | 1.000-1.246 | Yes |
| | | Socioeconomic status (PPP) | 1.000000 | 1.000000-1.000493 | Yes |
| HAALSI | KLoSA | Age | 1.073 | 1.049-1.098 | Yes |
| | | Female | 1.394 | 1.057-2.168 | Yes |
| | | Ever Smoked | 2.569 | 0.983-5.251 | No |
| | | Diabetes Treatment | 1.316 | 0.549-4.722 | No |
| | | Hypertension Medication | 1.808 | 1.018-4.815 | Yes |
| | | Medical Insurance | 0.878 | 0.488-1.278 | No |
| | | Socioeconomic status (PPP) | 1.001554 | 0.999996-1.004285 | No |
| SHARE | HRS+CHARLS+ KLoSA+LASI+MHAS | Age | 1.037 | 1.024-1.050 | Yes |
| | | Female | 2.339 | 1.890-2.925 | Yes |
| | | Ever Smoked | 2.269 | 1.000-5.524 | No |
| | | Diabetes Treatment | 0.696 | 0.486-0.996 | Yes |
| | | Hypertension Medication | 1.819 | 1.442-2.249 | Yes |
| | | Medical Insurance | 0.774 | 0.637-0.951 | Yes |
| | | Socioeconomic status (PPP) | 1.000004 | 0.999998-1.000013 | No |
| ELSA | HRS+CHARLS+ MHAS+HAALSI+SHARE | Age | 1.000 | 0.975-1.035 | No |
| | | Female | 1.042 | 0.685-1.912 | No |
| | | Ever Smoked | 0.851 | 0.423-1.216 | No |
| | | Diabetes Treatment | 0.489 | 0.267-1.000 | Yes |
| | | Hypertension Medication | 5.888 | 1.000-153.816 | Yes |
| | | Medical Insurance | 1.000 | 1.000-1.000 | Yes |
| | | Socioeconomic status (PPP) | 1.000000 | 0.998337-1.001234 | No |

Table S12: Inference results for the optimal transferable models in the HRS racial/ethnic groups.

| Target cohort | Source combination | Predictor | OR | 95% CI | Significant |
|---|---|---|---|---|---|
| Black | Hispanic+White | Age | 0.998 | 0.981-1.012 | No |
| | | Female | 1.042 | 0.815-1.339 | No |
| | | Education: GED | 1.030 | 0.574-1.653 | No |
| | | Education: HS Graduate | 0.709 | 0.464-0.988 | Yes |
| | | Education: Some College | 0.593 | 0.388-0.850 | Yes |
| | | Education: College+ | 0.338 | 0.184-0.552 | Yes |
| | | Wealth (per $100k) | 0.963 | 0.892-0.997 | Yes |
| | | Smoking: Former | 0.809 | 0.520-0.990 | Yes |
| | | Smoking: Never | 0.695 | 0.441-0.987 | Yes |
| | | BMI | 1.000 | 0.981-1.015 | No |
| | | Diabetes treatment: Oral only | 0.980 | 0.800-1.311 | No |
| | | Diabetes treatment: Any insulin | 1.329 | 1.037-2.035 | Yes |
| | | Medicare | 0.979 | 0.815-1.387 | No |
| | | Medicaid | 1.202 | 0.960-1.646 | No |
| | | Private insurance | 0.840 | 0.625-1.075 | No |
| Hispanic | White+Black | Age | 0.994 | 0.974-1.006 | No |
| | | Female | 1.208 | 1.002-1.668 | Yes |
| | | Education: GED | 0.745 | 0.354-1.129 | No |
| | | Education: HS Graduate | 0.739 | 0.490-0.991 | Yes |
| | | Education: Some College | 0.390 | 0.260-0.606 | Yes |
| | | Education: College+ | 0.351 | 0.164-0.646 | Yes |
| | | Wealth (per $100k) | 0.932 | 0.842-0.981 | Yes |
| | | Smoking: Former | 0.981 | 0.683-1.373 | No |
| | | Smoking: Never | 0.981 | 0.719-1.461 | No |
| | | BMI | 1.000 | 0.976-1.015 | No |
| | | Diabetes treatment: Oral only | 0.796 | 0.583-0.988 | Yes |
| | | Diabetes treatment: Any insulin | 1.188 | 0.890-1.801 | No |
| | | Medicare | 0.978 | 0.719-1.303 | No |
| | | Medicaid | 1.228 | 0.949-1.702 | No |
| | | Private insurance | 0.671 | 0.468-0.923 | Yes |
| Other | White+Black | Age | 0.999 | 0.999-1.046 | No |
| | | Female | 1.026 | 0.580-1.234 | No |
| | | Education: GED | 1.033 | 0.160-1.053 | No |
| | | Education: HS Graduate | 1.002 | 0.692-1.771 | No |
| | | Education: Some College | 0.982 | 0.539-1.308 | No |
| | | Education: College+ | 0.939 | 0.427-0.995 | Yes |
| | | Wealth (per $100k) | 0.998 | 0.982-1.018 | No |
| | | Smoking: Former | 0.992 | 0.917-1.861 | No |
| | | Smoking: Never | 0.972 | 0.568-1.153 | No |
| | | BMI | 1.000 | 0.970-1.030 | No |
| | | Diabetes treatment: Oral only | 0.990 | 0.831-1.773 | No |
| | | Diabetes treatment: Any insulin | 1.032 | 0.273-1.045 | No |
| | | Medicare | 1.246 | 0.993-2.824 | No |
| | | Medicaid | 1.228 | 1.030-3.308 | Yes |
| | | Private insurance | 0.777 | 0.366-0.985 | Yes |
| White | Hispanic+Black | Age | 1.008 | 0.994-1.025 | No |
| | | Female | 1.261 | 1.026-1.566 | Yes |
| | | Education: GED | 0.806 | 0.501-1.375 | No |
| | | Education: HS Graduate | 0.691 | 0.508-0.995 | Yes |
| | | Education: Some College | 0.554 | 0.398-0.941 | Yes |
| | | Education: College+ | 0.448 | 0.313-0.762 | Yes |
| | | Wealth (per $100k) | 0.991 | 0.971-1.000 | No |
| | | Smoking: Former | 0.530 | 0.374-0.987 | Yes |
| | | Smoking: Never | 0.471 | 0.333-0.877 | Yes |
| | | BMI | 0.997 | 0.980-1.012 | No |
| | | Diabetes treatment: Oral only | 0.960 | 0.753-1.225 | No |
| | | Diabetes treatment: Any insulin | 1.225 | 0.925-1.681 | No |
| | | Medicare | 0.774 | 0.533-1.024 | No |
| | | Medicaid | 1.441 | 1.100-2.136 | Yes |
| | | Private insurance | 0.745 | 0.580-0.936 | Yes |
